# The expression of insulin signaling and N-methyl-D-aspartate receptor genes in areas of gray matter atrophy is associated with cognitive function in type 2 diabetes

**DOI:** 10.1101/2025.03.26.25324696

**Authors:** Shelli R. Kesler, Heather Cuevas, Kimberly A. Lewis, Oscar Y. Franco-Rocha, Elena Flowers

**Author notes:** Corresponding Author: Shelli Kesler.

## Abstract

Type 2 diabetes (T2DM) is associated with brain abnormalities and cognitive dysfunction, including increased risk for Alzheimer’s disease. However, the mechanisms of T2DM-related dementia remain poorly understood. We obtained retrospective data from the Mayo Clinic Study of Aging for 271 individuals with T2DM and 542 demographically matched non-diabetic controls (age 51-89, 62% male). We identified regions of significant gray matter atrophy in the T2DM group and then determined which genes were significantly expressed in these brain regions using imaging transcriptomics. We selected 15 candidate genes involved in insulin signaling, lipid metabolism, amyloid processing, N-methyl-D-aspartate-mediated neurotransmission, and calcium signaling. The T2DM group demonstrated significant gray matter atrophy in regions of the default mode, frontal-parietal, and sensorimotor networks (p < 0.05 cluster threshold corrected for false discovery rate, FDR). *IRS1, AKT1, PPARG, PRKAG2*, and *GRIN2B* genes were significantly expressed in these same regions (R^2^ > 0.10, p < 0.03, FDR corrected). Bayesian network analysis indicated significant directional paths among all 5 genes as well as the Clinical Dementia Rating score. Directional paths among genes were significantly altered in the T2DM group (Structural Hamming Distance = 12, p = 0.004), with *PPARG* expression becoming more important in the context of T2DM-related pathophysiology. Alterations of brain transcriptome patterns occurred in the absence of significant cognitive deficit or amyloid accumulation, potentially representing an early biomarker of T2DM-related dementia.

## Introduction

Changes in brain structure and brain activation patterns are found in obesity and insulin resistance before development of overt type 2 diabetes mellitus (T2DM), which indicates that many who develop this condition may already have alterations in cognitive function (1, 2). Given these early changes, it is notable that T2DM, cognitive dysfunction, aging, and Alzheimer’s Disease (AD) share several common genetic and environmental risk factors (3–5). Overlapping molecular mechanisms include those involved in insulin signaling, lipid metabolism, inflammation, and amyloid processing (6–8). Many of these genes also play a role in neuronal protection, synaptic plasticity, neurodevelopment, and neurogenesis and may potentially interact with genes that regulate neurotransmission. Imaging transcriptomics reveals how gene expression varies across different regions of the brain, highlighting the molecular characteristics that underlie disease-related differences in brain structure and function between these regions (9–12). Transcriptomic brain atlases allow co-localization of neuroimaging and gene expression data to study in vivo molecular mechanisms of brain health and disease. Although single nucleotide polymorphism (SNP) studies have greatly enhanced our understanding of the genetic foundations of neurobiological traits and disorders, imaging transcriptomics provides a more detailed and functionally relevant perspective. By directly associating gene expression patterns with brain structure and function, imaging transcriptomics facilitates new opportunities for investigating the molecular mechanisms underlying neurological diseases and developing targeted therapeutic interventions.

While several imaging transcriptomics studies have been conducted in AD and other neurodegenerative disorders (13–15), very few have involved individuals with diabetes. Liu et al. (2025) found 298 genes that were co-localized with differences in brain morphology in individuals with T2DM and controls. Representative genes from gene clusters included *TSHZ1, CAMTA1, COG5,* and *SLC17A8* (16). Zhang and colleagues (2023) found that regions with disrupted functional connectivity in T2DM compared to controls were co-localized with two genetic components. These components were enriched for the neuronal system and potassium channels pathways, as well as biological processes such as neurotransmitter secretion, synaptic signaling, ion transmembrane transport, TrkB-Rac1 signaling pathway, and skeletal system morphogenesis/development (17).

These studies explored global gene expression patterns, but not specific pathways involved in diabetes-related neurodegeneration. Furthermore, gene-brain co-expression patterns were not linked to cognitive function. We selected a priori 10 candidate genes based on their critical roles in insulin signaling, lipid metabolism, and amyloid processing, which influence disease pathways such as impaired glucose utilization in the brain, chronic neuroinflammation, and accumulation of toxic protein aggregates. We also selected 5 candidate genes based on their roles in N-methyl-D-aspartate (NMDA)-mediated neurotransmission and calcium signaling, which support advanced cognitive functions, for a total of 15 genes (Table 1). We hypothesized that these genes would be expressed in regions of gray matter atrophy in individuals with T2DM and that they would interact to affect cognitive function.

**Table 1.** Genes selected for imaging transcriptomics analysis.

| Gene | Role | Relevance to Brain Function | References |
| --- | --- | --- | --- |
| *INSR* | Insulin receptor, initiates insulin signaling cascades | Synaptic plasticity, cognitive processing, and neuroprotection | (60-63) |
| *IRS1/2* | Insulin receptor substrate, mediates insulin signaling | Neuronal survival, synaptic function, and memory formation | (64-67) |
| *AKT1* | Key kinase in insulin signaling, regulates metabolism and neuroprotection | Synaptic maintenance, and neuroprotection | (68-71) |
| *IGF1* | Insulin-like growth factor, supports neurogenesis and glucose metabolism | Synaptic plasticity, cognition, and neurogenesis | (72-74) |
| *PPARG* | Regulates glucose metabolism, insulin sensitivity, and neuroprotection | Neuroinflammation, metabolic stress, and neuronal integrity | (51-54) |
| *SLC2A1/3*<br>(*GLUT1/3*) | Glucose transporter in endothelial cells, supports blood-brain barrier | Maintains glucose supply to the brain, essential for neuronal function | (8, 75-78) |
| *PRKAG2* | Insulin sensitivity, glucose metabolism, energy homeostasis | Neuronal metabolism and survival, autophagy regulation | (6, 7, 50) |
| *GRIN2A/B* | NMDA receptors | Neuronal development, excitatory neurotransmission, synaptic plasticity, neuroprotection | (42-46) |
| *CACNA1C* | Calcium signaling | Neurogenesis,<br>neurotransmitter<br>release, synaptic<br>plasticity, gene<br>expression | (44, 79-82) |
| *ATP2B4* | Calcium transport<br>and homeostasis | Intracellular calcium<br>levels, synaptic<br>function, protects<br>against excitotoxicity | (44, 79, 83, 84) |
| *ADCYAP1* | cAMP signaling,<br>stress response | Neuroprotection,<br>immune modulation,<br>inflammatory signaling | (44, 79, 85, 86) |
| *APOE* | Lipid metabolism,<br>transport, and<br>homeostasis | Synaptic remodeling,<br>amyloid clearance,<br>neuronal protection,<br>neuroinflammation | (87-90) |

## Methods

### Participants

We obtained data from the Mayo Clinic Study of Aging (MCSA) via the Laboratory of Neuroimaging’s Image and Data Archive at the University of Southern California. The MCSA is a prospective, epidemiologic study of cognitive aging that began in 2004 in Olmsted County, Minnesota (18). The Mayo Clinic and Olmsted Medical Center institutional review boards approved the parent study from which this data was extracted and gave approval for the sharing of anonymized data. All participants provided written informed consent during the parent study. For the current study, privacy rights of participants were observed, and no personally identifiable information was obtained. The Image and Data Archive manages and facilitates the de-identification, integration, visualization, and sharing of neuroscience data. It supports multi-site collaborations worldwide by providing a robust infrastructure for data preservation and exploration, while allowing study investigators to retain control over their data.

During the MCSA, participants underwent a medical history review, neurological evaluation, and provided demographic information. Trained nurses reviewed medical records to obtain data on chronic conditions including diagnosis of T2DM (19, 20). At the time of our study, there were N = 22,239 cases in the Image and Data Archive, but only 1,802 had magnetic resonance imaging (MRI) data and N=271 of these had T2DM. Missing data were assessed using Little’s MCAR test (21) and visualizations, which indicated that they were missing at random (MAR). We first excluded variables with greater than 5% missingness and then performed multiple imputation using a random forest model. We generated 10 imputed datasets, and all analyses were conducted on the pooled data following Rubin’s Rules to appropriately combine estimates, standard errors, and variance across imputations (22). Imputed data were evaluated using visualizations to ensure that imputed values were within a plausible range, followed the observed pattern, and had a similar distribution to observed data.

To create a comparable control group, we applied nearest neighbor propensity score matching. All cases with T2DM were retained, while a subset of non-diabetes control cases was selected using a 2:1 nearest neighbor matching algorithm based on a propensity score estimated from a logistic regression model. The propensity score was derived from key demographic variables, including age, ethnicity, race, sex, and education level. This approach ensures that each diabetes case was matched to a non-diabetic control case with the most similar propensity score, minimizing systematic differences between groups. Balance diagnostics were conducted using standardized mean differences, with a threshold of < 0.1 indicating adequate balance. The final dataset comprised all T2DM cases (N = 271) and their matched control counterparts (N = 542), allowing for direct group comparisons while reducing confounding effects (Figure 1).

**Figure 1.**
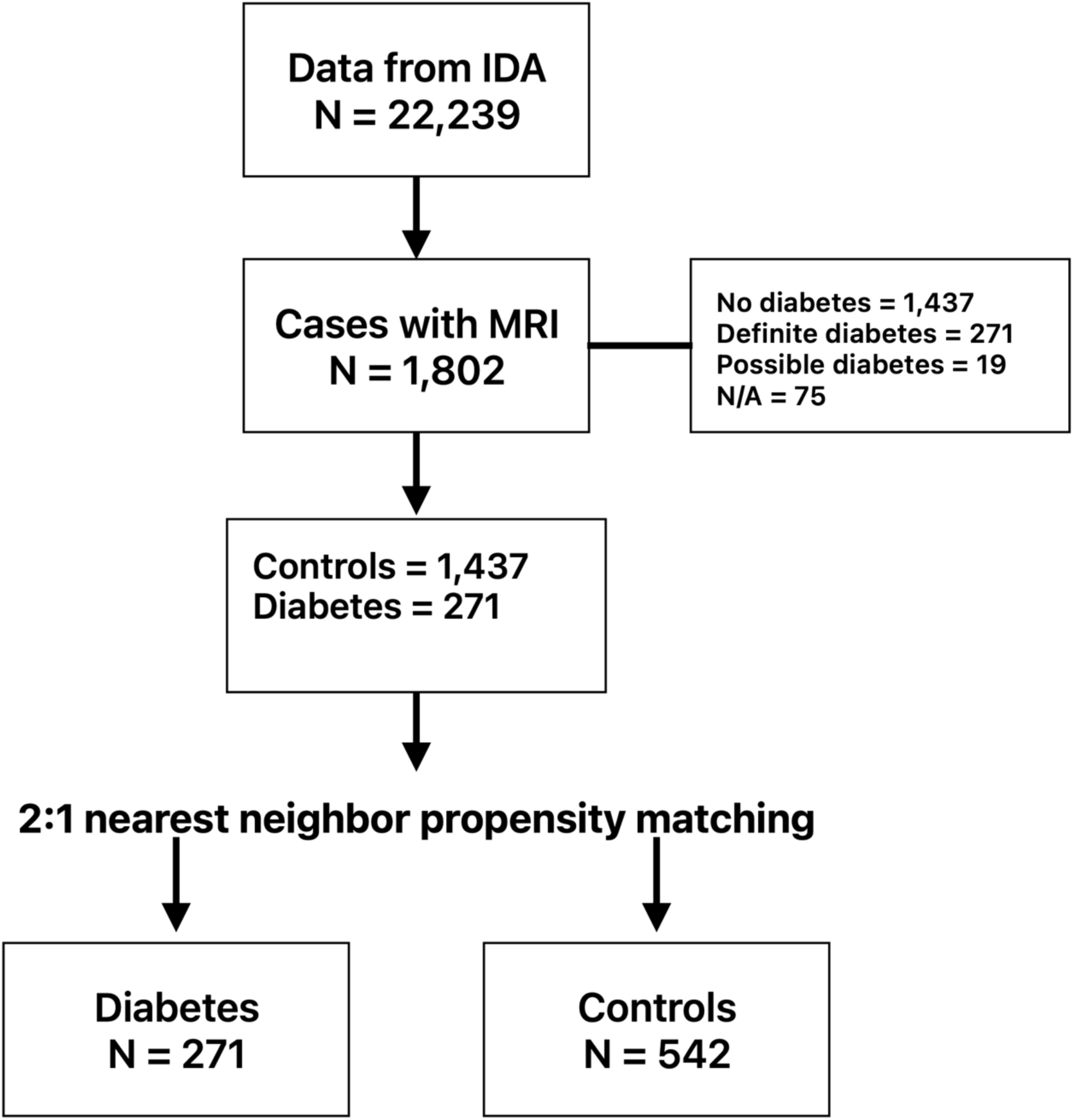
Sample selection. Of the N=22,239 cases in the Imaging Data Archive (IDA), 1,802 had available MRI data. Cases with “possible diabetes” or missing diabetes diagnostic data (N/A) were excluded. Propensity matching was conducted to identify a control group that was highly similar to the diabetes group in terms of sex, age, race, ethnicity and education.

### Cognitive Assessments

During the MCSA, participants were administered several neurological and neuropsychological assessments. For this study, we examined data from the Clinical Dementia Rating (CDR) scale, the Mini Mental State Exam, and the MCSA neuropsychological battery (18). The neuropsychological battery was administered by a trained psychometrist and included tests of memory (Auditory Verbal Learning Test Delayed Recall Trial, Wechsler Memory Scale-Revised Logical Memory II and Visual Reproduction II), language (Boston Naming Test, Category Fluency), executive function (Trail Making Test B, Wechsler Adult Intelligence Scale-Revised Digit Symbol, and visuospatial skills (Wechsler Adult Intelligence Scale-Revised Picture Completion and Block Design).

Raw scores for each neuropsychological test were normalized into age- and education-adjusted scores using Mayo’s Older American Normative Studies data. Cognitive domain scores were computed by adding the normalized scores of the tests included within each domain and scaled to allow comparisons across domains. A diagnosis of normal cognition, mild cognitive impairment (MCI), dementia or AD was made by consensus, considering all the data collected (history, neuropsychological testing, CDR scores, etc.) (18).

### Gray Matter Volume Measurement

Anatomic brain MRIs were performed on all consenting individuals with no contraindications. Those who underwent MRI were not significantly different demographically from those who did not, per previous report (23). Imaging data were collected with a 3 Tesla GE scanner (GE Medical Systems, Milwaukee, WI) using a 3D magnetization prepared rapid acquisition gradient echo (MPRAGE) sequence (repetition time = 2300 ms, echo time = 3 ms, inversion time = 900 ms, flip angle = 8°, field of view = 26 cm, in-plane matrix = 256 × 256, phase field of view = 0.94, slice thickness = 1.2 mm).

We extracted gray matter volumes from anatomic brain MRI using voxel-based morphometry implemented in Statistical Parametric Mapping (SPM) 12 (Wellcome Trust Centre for Neuroimaging, London, UK) (24). We employed Diffeomorphic Anatomical Registration Through Exponentiated Lie Algebra, which uses a large deformation framework to preserve topology and employs customized, sample-specific templates resulting in superior image registration. Successful segmentation and normalization were confirmed using visual and quantitative quality assurance methods. Regional differences between groups were measured using voxel-wise analyses with a general linear model framework to obtain probability maps indicating voxels characterized as gray matter. A two-sample t-test model was employed to create an SPM T-score contrast map for group comparisons (controls > T2DM; T2DM > controls) with a height threshold of p < 0.001 and a cluster threshold of p < 0.05 corrected for multiple comparisons using the false discovery rate (FDR) (25).

### Imaging Transcriptomics

We obtained brain transcriptome data from the Allen Human Brain Atlas (AHBA), which is currently considered the most comprehensive transcriptional brain map available (9). AHBA was developed using six adult human donor brains to provide expression data from tens of thousands of genes measured from thousands of brain regions (26). To determine the co-localization of brain imaging and transcription data, we used the Multimodal Environment for Neuroimaging and Genomic Analysis toolbox v3.1 (27) to correlate the control > T2DM SPM contrast map with patterns of AHBA expression from the 15 selected genes (T2DM > controls map was not significant). The SPM contrast map was resampled into AHBA coordinates with a 5mm resolution. The expression data for each AHBA donor, for each gene was sampled from each of 169 AHBA regions resulting in a 169×6 region-by-gene expression matrix for each gene. Principal Components Analysis was performed on the region-by-gene expression matrix to identify components explaining at least 95% variance across the AHBA donors. The component scores were then entered as independent variables into a multiple linear regression analysis with the corresponding gray matter volume values from the SPM contrast map as the dependent variable. P values for the regression models were corrected for multiple comparisons using FDR. We also fit a first-degree polynomial of the regression model, extracting the coefficients for both the slope and intercept. To determine the direction of the relationship between gene expression and gray matter volume, we calculated the sign of the slope coefficient using the sign function.

Many AHBA genes have multiple probes with variable reliability in terms of expression patterns. Therefore, the accuracy of the probe to gene mapping was verified and probe data were normalized to z-scores. Representative probes were then selected in a data-driven manner considering between-donor homogeneity and the distributions of probe data. Genomic autocorrelations were calculated using an intraclass correlation coefficient (ICC) to measure the gene expression variability between donors.

### APOE Genotype and Amyloid PET Imaging

A 10-hour fasting blood draw was performed with each participant from which DNA was extracted, including apolipoprotein E (*APOE*) genotype. Participants were classified as having presence or absence of the AD risk-related *APOE4* alleles (18). Amyloid positron emission tomography (PET) imaging was performed with Pittsburgh Compound B to measure amyloid deposition in key cortical regions. A standardized uptake value ratio (SUVR) was computed from the voxel-number weighted average of the median uptake in prefrontal, orbitofrontal, parietal, temporal, anterior and posterior cingulate, and precuneus regions normalized to the cerebellar crus gray median. Elevated amyloid PET was defined as a SUVR value greater than or equal to 1.48 (28).

### Statistical Analyses

Demographic, medical, cognitive, *APOE* genotype and amyloid PET data were compared between groups using two-sample, two-tailed Wilcoxon tests for continuous variables and Chi squared tests for categorical variables (p < 0.05). R squared values for the genes that demonstrated significant co-localization with the control > T2DM SPM contrast map (i.e., genes that were significantly expressed in regions of gray matter atrophy) were extracted for each participant. These values represent the strength of the image-transcriptome co-localization. R squared values were correlated with medication status (insulin, metformin, blood pressure lowering medication, statins) in the T2DM group using Spearman rho (two-tailed, p < 0.05, FDR corrected).

R squared values were also entered into a Bayesian network analysis to examine the causal relationships among significant genes and cognitive measures. Bayesian network analysis identifies conditional dependencies between variables without requiring prior assumptions about their relationships (29). The optimal network structure was determined using a hill-climbing algorithm, which maximized the Bayesian Information Criterion (BIC) to balance model complexity and goodness of fit (30). The resulting conditional relationships among variables were represented as directional edges in a directed acyclic graph (DAG) (31). A Bayesian network was determined for each group and the networks were compared using the Structural Hamming Distance (SHD) (32). SHD measures how many edge modifications are needed to transform one network into another. These modifications include edge addition (adding an edge that exists in one network but not in the other), edge deletion (removing an edge that exists in one network but not in the other), and edge reversal (reversing the direction of an edge). The SHD was assessed for significance using permutation testing. A total of 5,000 random DAGs were generated, each maintaining the same edge probabilities as the original networks. These random DAGs were used to construct a permutation distribution of SHD values. The p-value was then calculated as the proportion of instances where the SHD from the random permutations exceeded the actual SHD.

## Results

Compared to controls, the T2DM group showed significantly higher CDR scores, including both the Sum of Boxes (W = 68193, p = 0.007) and Final Score (W = 68617, p = 0.010). The two groups did not differ significantly in terms of cognitive impairment status, Mini Mental Status Exam scores, or performance on the MCSA neuropsychological battery (Table 2).

**Table 2.**
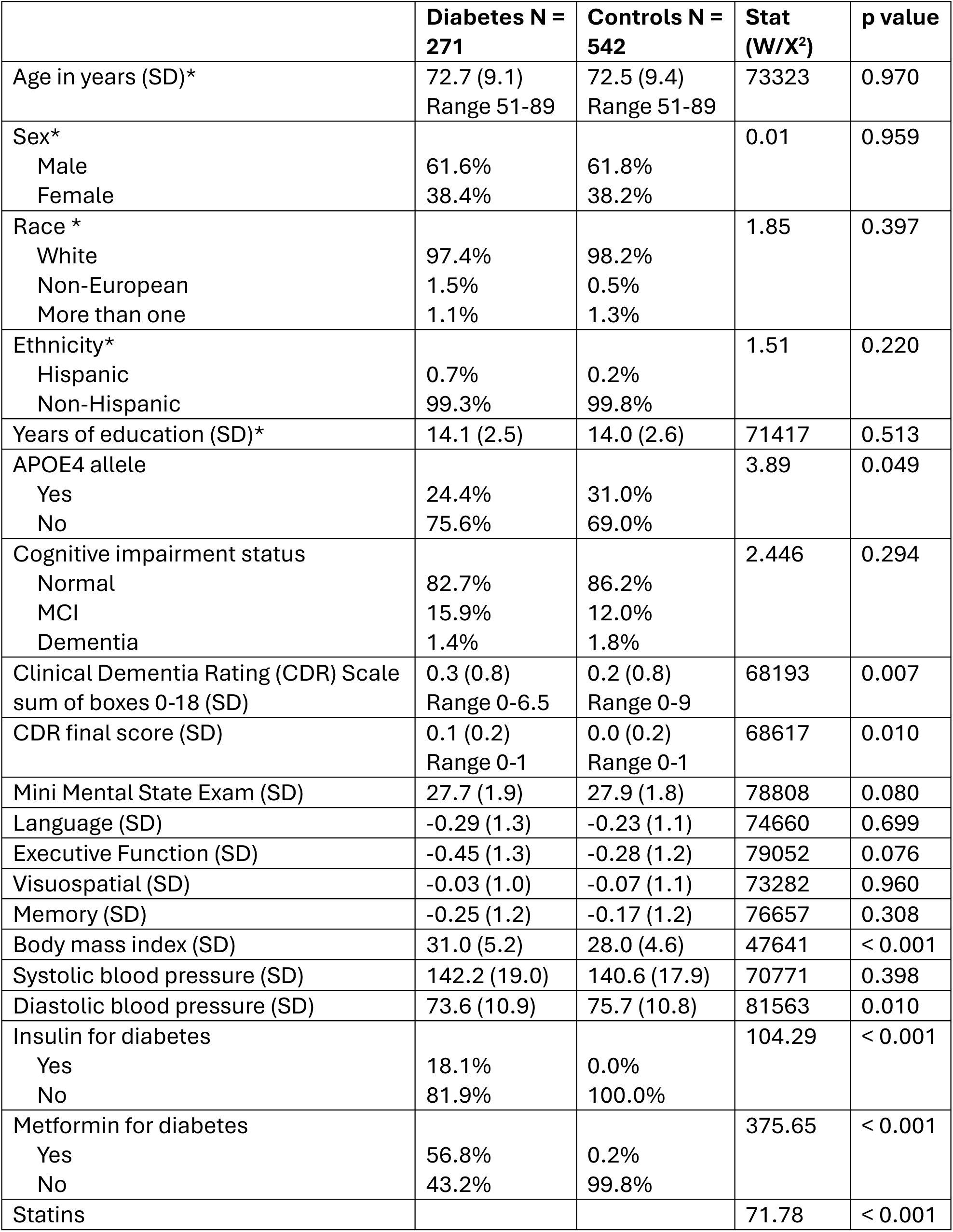
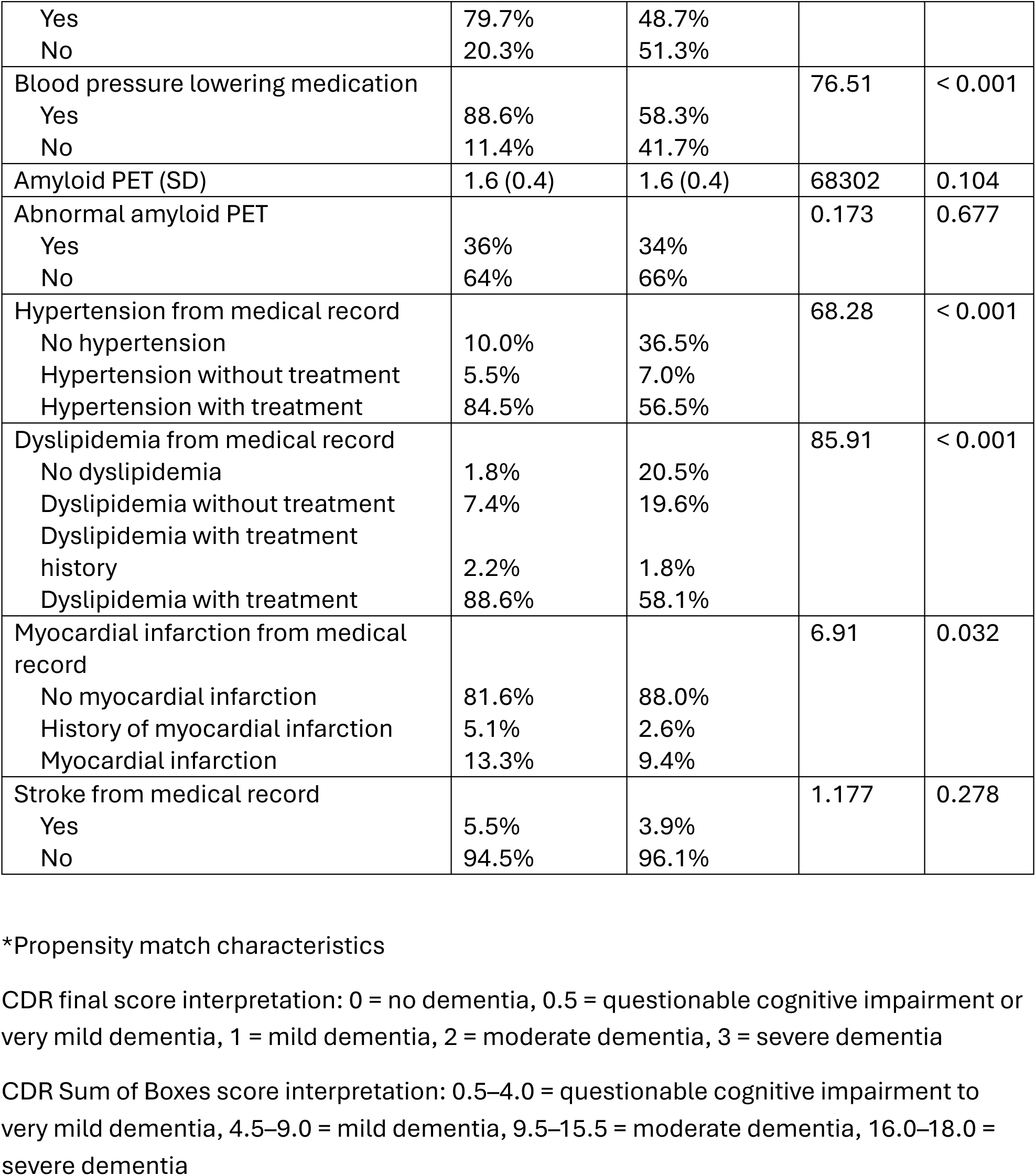
Demographic and clinical characteristics per group. SD = standard deviation.

VBM analysis indicated significant, widespread gray matter atrophy in the T2DM group compared to controls. Affected regions included bilateral frontal cortex, anterior and posterior cingulate, bilateral precuneus and postcentral gyrus, bilateral temporal cortex, bilateral thalamus, caudate, and putamen, bilateral calcarine cortex and lateral occipital cortex (Figure 2). The T2DM group did not demonstrate any regions of greater gray matter volume compared to controls.

**Figure 2.**
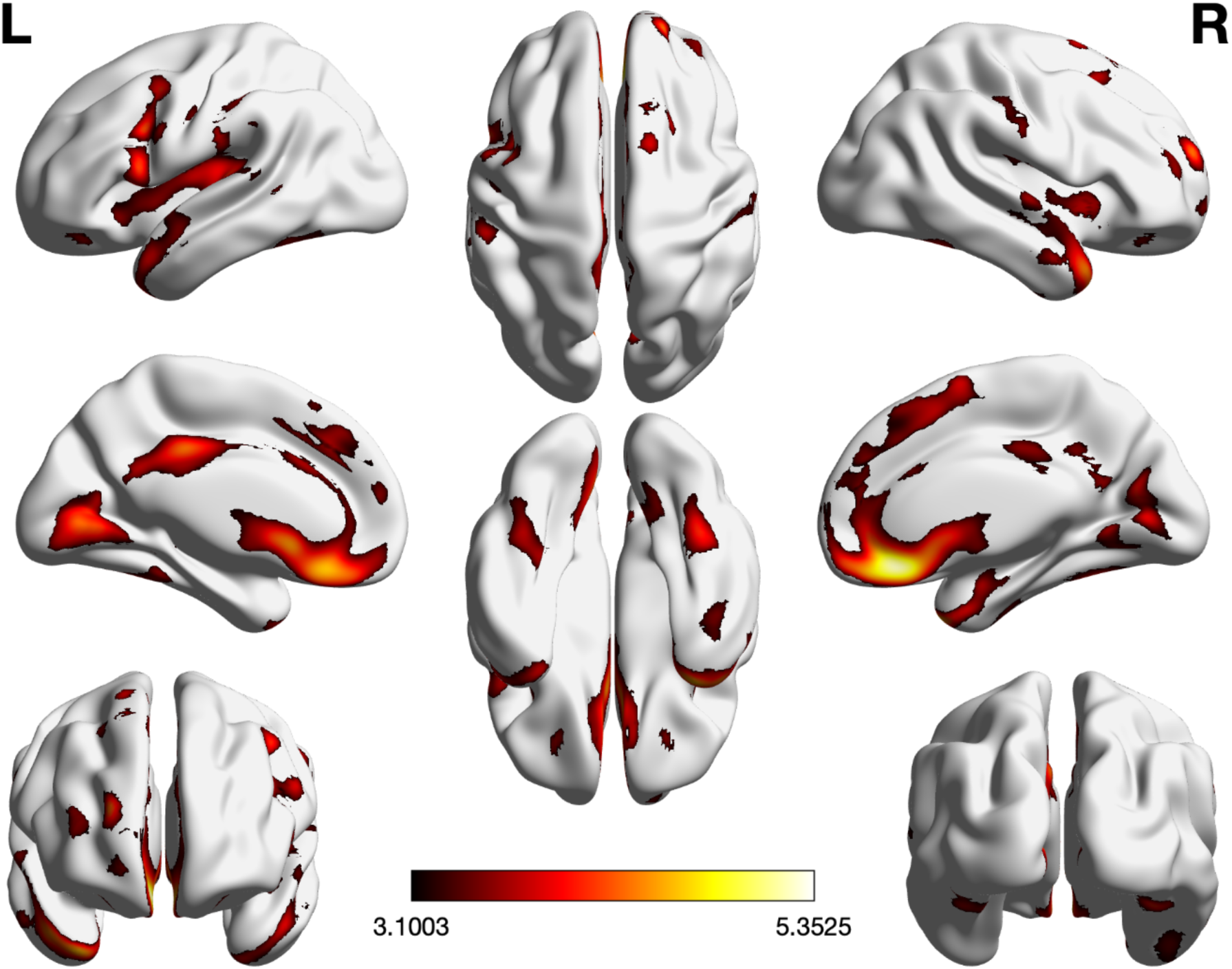
Voxel-based morphometry. Results indicated significant gray matter atrophy in the T2DM group compared to controls (p < 0.001 height, p < 0.05 FDR corrected cluster extent threshold). Affected regions included bilateral frontal cortex, anterior and posterior cingulate, bilateral precuneus and postcentral gyrus, bilateral temporal cortex, bilateral thalamus, caudate, and putamen, bilateral calcarine cortex and lateral occipital cortex. Color bar indicates the cluster T-score.

Imaging transcriptomics analysis demonstrated that spatial expression patterns for 5 genes were significantly correlated with gray matter volume in regions of atrophy in the T2DM group. In other words, gray matter volume of the regions shown in Figure 2 were significantly correlated with the expression patterns of *IRS1, AKT1, PPARG, PRKAG2*, and *GRIN2B* in those same regions (Table 3). *IRS1, AKT1, PPARG* showed direct correlations with gray matter volume while *PRKAG2* and *GRIN2B* showed inverse relationships.

**Table 3.** Imaging transcriptomics analysis results. The R squared value indicates the relationship between gene expression and gray matter volume in regions demonstrating atrophy in the T2DM group. D indicates the direction of this relationship based on the sign of the polynomial regression fit. FDR is the false discovery rate correction for multiple comparisons. ICC is the mean (**µ**) and standard deviation (**σ**) intraclass correlation coefficient for gene expression among Allen Human Brain Atlas donors. The ICC indicates the reliability of gene expression across donors.

| | R<sup>2</sup> | D | F | p | FDR p | ICCμ | ICCσ |
| --- | --- | --- | --- | --- | --- | --- | --- |
| *INSR* | 0.129 | 1 | 2.467 | **0.039** | 0.098 | 0.533 | 0.101 |
| *IRS1* | 0.158 | 1 | 3.934 | **0.006** | **0.024** | 0.842 | 0.036 |
| *IRS2* | 0.123 | -1 | 2.322 | **0.050** | 0.108 | 0.726 | 0.091 |
| *AKT1* | 0.174 | 1 | 3.506 | **0.006** | **0.024** | 0.642 | 0.168 |
| *IGF1* | 0.050 | -1 | 0.879 | 0.499 | 0.576 | 0.701 | 0.094 |
| *PPARG* | 0.248 | 1 | 5.477 | **2.09E-04** | **0.003** | 0.654 | 0.101 |
| *PRKAG2* | 0.158 | -1 | 3.943 | **0.006** | **0.024** | 0.853 | 0.055 |
| *SLC2A1* | 0.061 | 1 | 1.084 | 0.375 | 0.469 | 0.710 | 0.133 |
| *SLC2A3* | 0.027 | 1 | 0.384 | 0.887 | 0.887 | 0.338 | 0.196 |
| *GRIN2A* | 0.041 | -1 | 1.834 | 0.166 | 0.226 | 0.941 | 0.014 |
| *GRIN2B* | 0.100 | -1 | 4.757 | **0.011** | **0.033** | 0.956 | 0.013 |
| *CACNA1C* | 0.091 | -1 | 1.654 | 0.155 | 0.226 | 0.829 | 0.043 |
| *ATP2B4* | 0.069 | -1 | 2.091 | 0.107 | 0.201 | 0.913 | 0.024 |
| *ADCYAP1* | 0.009 | 1 | 0.395 | 0.675 | 0.723 | 0.939 | 0.010 |
| *APOE* | 0.089 | -1 | 1.614 | 0.165 | 0.226 | 0.680 | 0.124 |

Significantly more participants in the T2DM group took insulin, metformin, blood pressure lowering medication, and statins compared to controls (p < 0.001, Table 2). After FDR correction, strength of *PRKAG2* co-localization with gray matter volume was significantly positively correlated with metformin (r = 0.25, p < 0.001) and negatively correlated with blood pressure lowering medications (r = −0.17, p = 0.022). No other correlations between image-gene co-localization and medications were significant.

There was a 7% higher incidence of *APOE4* genotypes in the control group which was moderately significant (X^2^ = 3.89, p = 0.049). However, the groups did not differ in terms of amyloid PET uptake (W = 68302, p = 0.104) or abnormal amyloid PET incidence (X^2^ = 0.173, p = 0.677, Table 2).

Bayesian network analysis (Figure 3) indicated significant directional paths (i.e., acyclic graph edges) among *IRS1, AKT1, PPARG, PRKAG2*, and *GRIN2B* and CDR. We evaluated the CDR Sum of Boxes score given that it was the most significant between groups. Both groups showed upregulation of *PRKAG2* by *GRIN2B* with higher expression of *PRKAG2* resulting in lower CDR scores. *AKT1* and *GRIN2B* were upregulated by *IRS1* in both groups. *IRS1* upregulated *PRKAG2* in the T2DM group but downregulated it in controls. *IRS1* and *GRIN2B* downregulated *PPARG* in controls but these relationships were missing in the T2DM group (i.e., false negative edges). *PRKAG2* and *AKT1* upregulated *PPARG* in controls but these paths were reversed in the T2DM group (i.e., false positive edges). The differences between the two networks were significant (SHD = 12, 95%CI = 4 to 11, p = 0.004; note that for permutation testing, significance is associated with a value outside the CI range). Given the significant Spearman rho correlation between metformin and *PRKAG2* as well as between blood pressure lowering medication and *PRKAG2*, we conducted a post-hoc Bayesian network in the diabetes group, which showed no significant edges for these medication variables.

**Figure 3.**
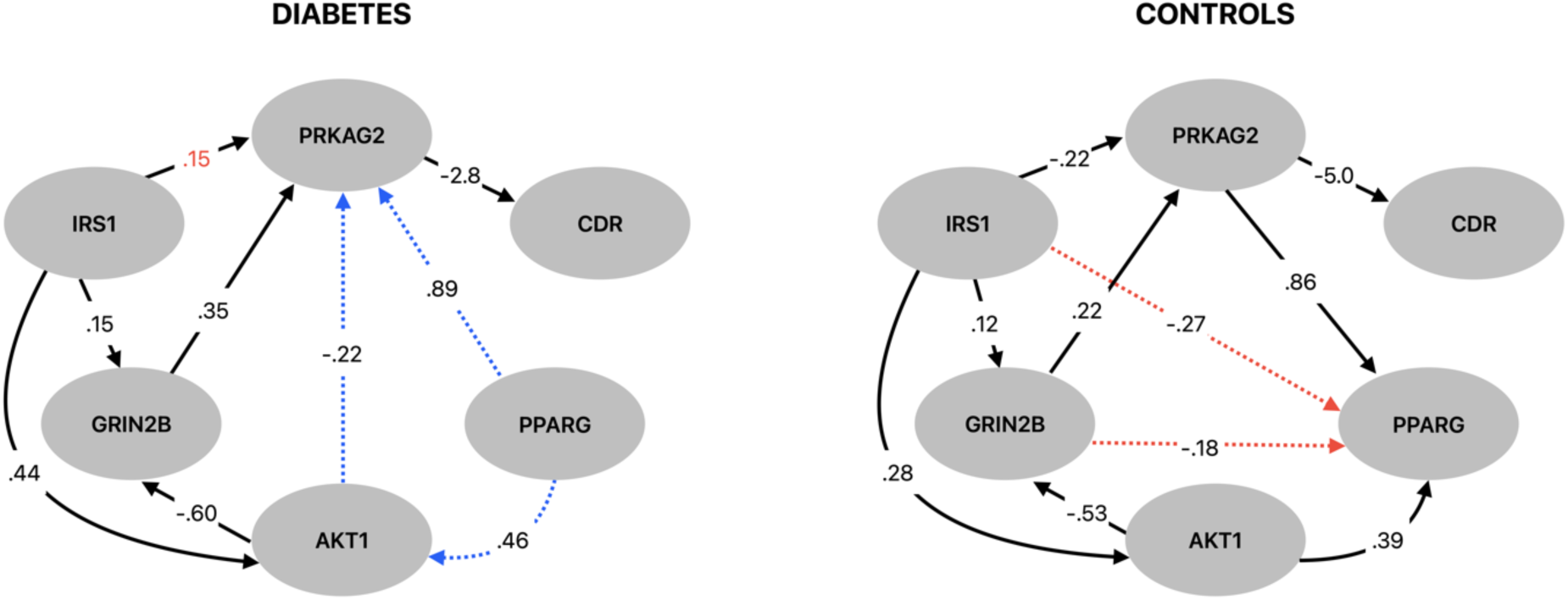
Bayesian network analysis indicated significant directional paths (i.e., direct acyclic graph edges) among genes whose expression patterns were co-localized with regions of gray matter atrophy in the diabetes group and Clinical Depression Rating (CDR) scale Sum of Boxes score. Black lines indicate common paths between groups in terms of existence and directionality. Blue lines indicate false positive paths in the T2DM group in terms of existence or directionality. Red lines signify false negative paths in the T2DM group in terms of existence or directionality. Red coefficients are those that have reversed sign in the T2DM group. Standardized coefficients are shown.

## Discussion

We evaluated gray matter morphology, cognitive function, and gene expression in individuals with T2DM compared to matched, non-diabetic controls. CDR scores were significantly elevated in the T2DM group compared to controls. However, these scores did not suggest MCI or dementia, on average, based on score interpretation guidelines (33). Both groups had a small percentage of individuals classified as having mild dementia (i.e., CDR scores >= 4.5). While not clinically significant, the elevation in CDR scores among T2DM individuals may represent an accelerated aging process that increases risk for cognitive dysfunction. Performance on standardized neuropsychological tests was equivocal between groups and was, on average, within the normal range (i.e., less than 0.5 standard deviations from the mean). Controls had a slightly higher incidence of *APOE4* genotype, but the groups did not differ in terms of cortical amyloid burden.

However, despite relatively equivalent neuropsychological performance, amyloid deposition, and incidence of *APOE4* genotype, there was significant gray matter atrophy affecting all cerebral lobes and several subcortical structures in the T2DM group. The affected regions are known to be involved in the default mode network (DMN), prefrontal-parietal network, and motor/sensory networks. These networks support higher cognitive function, sensory-motor integration, and emotional regulation. Our findings were consistent with those from prior studies of T2DM (34–37). The mechanisms of gray matter atrophy in T2DM are likely multifactorial. The brain is the largest consumer of metabolic resources compared to other organs and T2DM involves disruption of glucose regulation. Regions in the DMN exhibit the highest glucose metabolic rates, (38) making them especially susceptible to disruptions in metabolism. The DMN is also the initial site of amyloid accumulation in the brain (39). Regions in the DMN and prefrontal networks regions are highly susceptible to the chronic stress and inflammation that often characterize T2DM. Prefrontal cortex functions rely on NMDA-mediated neurotransmission and calcium signaling. Excessive activation of these pathways is associated with neuroinflammation and has been shown to play a role in neurodegenerative diseases (40–42).

Imaging transcriptomics analysis indicated that regions of atrophy in the T2DM group significantly expressed genes involved in NMDA-mediated neurotransmission (*GRIN2B*) and insulin signaling (*IRS1, AKT1, PPARG*, *PRKAG2*). *GRIN2B* encodes the NMDA receptor GluN2B subunit (also known as NR2B), which is involved in excitatory neurotransmission, synaptic plasticity, and neuroprotection (42–46). SNPs in *GRIN2B* have been associated with T2DM, as well as several T2DM-related complications, including cholesterol level, body mass index, retinopathy, hypertension, and cerebrovascular disease (47). GluN2B-containing NMDA receptors are involved in neuronal toxicity and cell death, as excessive calcium influx mediated by these receptors can result in cellular damage and apoptosis (48, 49). Accordingly, our findings indicated that gray matter atrophy in T2DM was associated with higher *GRIN2B* expression.

However, Bayesian network analysis indicated that *GRIN2B* upregulated *PRKAG2,* which in turn improved cognitive function, yet higher *PRKAG2* expression was correlated with gray matter atrophy. These findings highlight the importance of considering biological mechanisms in context. All genes included in the Bayesian network indirectly or directly affected cognitive function, although *PRKAG2* was the only gene that had a direct path to CDR score. *PRKAG2* is involved in neuronal metabolism and survival as well as autophagy regulation (6, 7, 50). The directional relationships among the genes were altered in the T2DM group with false positive and false negative paths originating from *PPARG*. This gene accounted for most of the variance in gray matter volume and may therefore represent a critical factor for diabetes-related brain morphology and cognitive function. *PPARG* has been shown to be involved in neuroinflammation, metabolic stress, and neuronal integrity (51–54). Interestingly, *PPARG* did not demonstrate a direct or indirect path to CDR in controls but exerted indirect effects via *AKT1* and *PRKAG2* due to edge reversals (i.e., path directionality reversals) in T2DM.

Our findings are consistent with studies supporting *PPARG* agonists as treatments for cognitive decline in both T2DM and AD (55). *PPARG* agonists, used in patients with T2DM and cognitive impairment, yield measurable benefits in both metabolic and cognitive domains. In controlled trials, adding rosiglitazone to metformin produced significant improvements in fasting plasma glucose and glycated hemoglobin (P < 0.05) and maintained cognitive performance on tests such as the Mini-Mental State Exam, Rey Auditory Verbal Learning Test, and Trail Making Tests in older adults with mild cognitive impairment (56). When used as an add-on therapy, rosiglitazone lowered working memory errors by 25–31% (P < 0.001 and P = 0.017, respectively) among older adults with T2DM (57).

Metformin usage was positively correlated with *PRKAG2* expression in gray matter while blood pressure lowering medications were negatively associated with *PRKAG2* expression. However, these medications did not have significant edges in a post-hoc Bayesian network for the T2DM group. Bayesian networks represent conditional dependencies rather than simple pairwise correlations. Thus, the relationship between medication usage and *PKARG2* may be fully mediated by the other genes in the model. Because the Bayesian network algorithm utilizes regularization to reduce overfitting, including medication effects may have increased model complexity without enough gain in explanatory power (i.e., the relationship was relatively weaker compared to other paths). Alternatively, unmodeled confounders may explain the relationships or the sample size may have lacked sufficient power for these additional variables.

Emerging evidence suggests that micro ribonucleic acids (miRNAs) acting as post-transcriptional regulators in the brain may further modulate the insulin signaling and metabolic pathways implicated in our findings (Table 4). In prior work examining metformin-responsive circulating miRNAs, we identified several miRNAs with potential relevance to the mRNA targets highlighted here (58). Specifically, metformin-associated downregulation of let-7c-5p and miR-222-3p may relieve repression of IRS2 and attenuate PTEN-mediated inhibition of AKT1, respectively, amplifying insulin signaling cascades. Additionally, the upregulation of miR-29b-3p and miR-93-5p intersects pathways governing glucose metabolism, autophagy, and synaptic resilience, aligning with the central role of *PRKAG2* and *AKT1* in our Bayesian network. These findings suggest that metformin’s effects may extend beyond glycemic control, influencing brain insulin signaling and neuronal metabolism through epigenetic mechanisms relevant to T2DM-related cognitive decline.

**Table 4.** Metformin-Responsive Circulating microRNA and Brain Insulin Signaling mRNA Pathways Mapping Table.

| **miRNA**<br>**(Metformin-**<br>**Responsive)** | **Direction of**<br>**Change** | **Known Targets /**<br>**Pathways** | **Relevance to mRNA**<br>**Targets in Current**<br>**Study** |
| --- | --- | --- | --- |
| let-7c-5p | ↓ Downregulated | INSR, IRS2,<br>ERBB4, NRG4,<br>DICER1 | IRS2 is upstream of<br>IRS1/AKT1 signaling<br>cascade |
| miR-222-3p | ↓ Downregulated | PTEN, MGMT,<br>PPP2R2A, RECK | PTEN regulates<br>PI3K/AKT1 pathway |
| miR-29b-3p | ↑ Upregulated | PER3, H19, MMP-2/9, fibrosis,<br>insulin<br>transcription,<br>autophagy | Links to<br>AKT/mTOR/autophagy<br>pathway |
| miR-93-5p | ↑ Upregulated | GLUT4, ERBB4,<br>NRG4 | Regulates glucose<br>metabolism<br>intersecting insulin<br>signaling |
| miR-130b-3p | ↓ Downregulated | PDHA1, GCK, lipid<br>metabolism, β-<br>cell ATP dynamics | Possible relevance to<br>metabolic regulation<br>and PPARG |

The limitations of this study include the retrospective, cross-sectional nature of the data. Although MCSA is a prospective, longitudinal study, MRI data were available only for a single timepoint. Most participants in our sample were White, male, and over the age of 65 years, reducing generalizability of our findings. We may have lacked power to detect significant expression of certain genes in regions of gray matter atrophy. The ICC for *SLC2A3* was low indicating relatively poor expression reliability across AHBA donors. Our selection of candidate genes was hypothesis driven; based on our domain knowledge, thorough review of the relevant literature, and consideration of our sample size, but other genes may be more important for brain morphology and cognitive function. The AHBA atlas includes expression patterns for thousands of brain regions but does not cover the entire brain. Different neuropsychological tests or cognitive assessments may yield larger between group effects. Even when data are MAR and imputed data are evaluated for quality and consistency, imputed values are estimates, introducing variability and potential model dependency that may affect downstream analyses.

Our findings contribute unique insights regarding the molecular mechanisms of cognitive dysfunction in T2DM. Specifically, insulin signaling and NMDA-mediated neurotransmission seem to be linked. *PPARG* may become more important under T2DM-related pathological conditions and *PPARG* agonists may influence cognitive function indirectly while *PRKAG2* may have a more direct role. We showed that alterations of brain transcriptome patterns occurred in the absence of significant cognitive deficit or amyloid accumulation, possibly representing an early biomarker of T2DM-related dementia. Future studies should examine whether the altered Bayesian network patterns demonstrated here could be linked to specific epigenetic markers (e.g., increased DNA methylation of a promoter region or altered histone acetylation). In addition to gray matter atrophy, T2DM has also been associated with abnormalities in functional connectivity and white matter integrity (59). Multimodal studies are needed to comprehensively evaluate imaging transcriptomic results.

## Acknowledgements

The data contained in this analysis were obtained under one of the following research grants from the National Institutes of Health to the Mayo Clinic Study of Aging (U01 AG06786, Ronald Petersen, PI) or the Mayo Alzheimer’s Disease Research Center (P30 AG062677, Ronald Petersen, PI).

## Data Availability

All data used in this study are available to qualified researchers and industry partners from the Laboratory of Neuroimaging’s Image and Data Archive at the University of Southern California or the Global Alzheimer’s Association Interactive Network.

---

**Supplementary Table 1.** Metformin-Responsive Circulating microRNA and Brain Insulin Signaling mRNA Pathways Mapping Table.

| **miRNA**<br>**(Metformin-Responsive)** | **Direction of Change** | **Known Targets / Pathways** | **Relevance to mRNA Targets in Current Study** |
| --- | --- | --- | --- |
| let-7c-5p | ↓ Downregulated | INSR, IRS2, ERBB4, NRG4, DICER1 | IRS2 is upstream of IRS1/AKT1 signaling cascade |
| miR-222-3p | ↓ Downregulated | PTEN, MGMT, PPP2R2A, RECK | PTEN regulates PI3K/AKT1 pathway |
| miR-29b-3p | ↑ Upregulated | PER3, H19, MMP-2/9, fibrosis, insulin transcription, autophagy | Links to AKT/mTOR/autophagy pathway |
| miR-93-5p | ↑ Upregulated | GLUT4, ERBB4, NRG4 | Regulates glucose metabolism intersecting insulin signaling |
| miR-130b-3p | ↓ Downregulated | PDHA1, GCK, lipid metabolism, β-cell ATP dynamics | Possible relevance to metabolic regulation and PPARG |

